# Comorbid Diabetes Mellitus was Associated with Poorer Prognosis in Patients with COVID-19: A Retrospective Cohort Study

**DOI:** 10.1101/2020.03.24.20042358

**Authors:** Yan Zhang, Yanhui Cui, Minxue Shen, Jianchu Zhang, Ben Liu, Minhui Dai, Linli Chen, Duoduo Han, Yifei Fan, Yanjun Zeng, Wen Li, Fengyu Lin, Sha Li, Xiang Chen, Pinhua Pan

## Abstract

**Background:** The 2019 novel coronavirus disease (COVID-19) emerged in Wuhan, Hubei province, China, and was characterized as pandemic by the World Health Organization. Diabetes mellitus is an established risk factor for poor clinical outcomes, but the association of diabetes with the prognosis of COVID-19 have not been reported yet.

**Methods:** In this cohort study, we retrospectively reviewed 258 consecutive hospitalized COVID-19 patients with or without diabetes at the West Court of Union Hospital of Huazhong University of Science and Technology in Wuhan, China, recruited from January 29 to February 12, 2020. The cases were confirmed by real-time PCR and the demographic, clinical, laboratory, radiological, and treatment data were collected and analyzed. Prognosis was defined as hospitalization, discharged survivor and death, which was followed up until March 12, 2020.

**Results:** Of the 258 hospitalized patients (63 with diabetes) with COVID-19, the median age was 64 years (range 23-91), and 138 (53.5%) were male. No significant differences in age and sex were identified between patients with and without diabetes. Common symptoms included fever (82.2%), dry cough (67.1%), polypnea (48.1%), and fatigue (38%). Patients with diabetes had significantly higher leucocyte and neutrophil counts, and higher levels of fasting blood glucose, serum creatinine, urea nitrogen and creatine kinase isoenzyme MB at admission compared with those without diabetes. COVID-19 patients with diabetes were more likely to develop severe or critical disease condition with more complications at presentation, and had higher incidence rates of antibiotic therapy, non-invasive and invasive mechanical ventilation, and death (11.1% vs. 4.1%). Cox proportional hazard model showed that diabetes (adjusted hazard ratio [aHR]=3.64; 95% confidence interval [CI]: 1.09, 12.21) and fasting blood glucose (aHR=1.19; 95% CI: 1.08, 1.31) were associated with the fatality of COVID-19, adjusting for potential confounders.

**Conclusions:** Diabetes mellitus is associated with greater disease severity and a higher risk of mortality in patients with COVID-19. Primary and secondary prevention strategies are needed for COVID-19 patients with diabetes.

## Introduction

Since December 2019, China has been experiencing an outbreak of pneumonia with a novel coronavirus [1], which was officially named as Severe Acute Respiratory Syndrome Coronavirus-2 (SARS-CoV-2) by the International Committee on Taxonomy of Viruses (ICTV). The World Health Organization (WHO) has declared that Coronavirus Disease 2019 (COVID-19) is caused by SARS-CoV-2 infection. WHO characterized COVID-19 as pandemic, as it has spread rapidly throughout China and more than 100 countries in the following two months after outbreak, causing more than 150,000 confirmed cases and thousands of death by March 15, 2020. SARS-CoV-2 belongs to the subgenus Sarbecovirus (β-CoV lineage B), and shares 79% of sequence with Severe Acute Respiratory Syndrome Coronavirus (SARS-CoV), but only 50% homology with Middle East Respiratory Syndrome Coronavirus (MERS-CoV) [2, 3].

China has a climbing prevalence rate of diabetes in recent decades. According to the latest nationally representative cross-sectional survey among 170287 participants in 2013 in mainland China, the overall prevalence of diabetes was 10.9% in adults and 20.2% in the elderly [4]. Diabetic patients are more susceptible to be infected by bacteria, virus, and fungus than individuals without diabetes owing to relatively lower immune function [5, 6]. As a result, these patients might be at an increased risk of SARS-CoV-2 infection and poorer prognosis.

Despite many studies have described the clinical characteristics of COVID-19 so far [1, 7, 8, 9], information with respect to diabetes among these patients, have not been well characterized yet. In the current study, we retrospectively reviewed the clinical data of 258 patients with laboratory-confirmed COVID-19, and compared the differences in clinical characteristics, laboratory markers, treatment strategies, and short-term prognosis including death between patients with and without diabetes. We hope that these findings will provide new insights into the risk stratification, disease management, and therapeutic strategies for diabetic patients with COVID-19.

## Methods

### Study design and participants

This was a retrospective cohort study among patients with COVID-19. All consecutive patients with laboratory-confirmed SARS-CoV-2 infection who were admitted to several isolation wards at the West Court of Union Hospital of Huazhong University of Science and Technology from January 29 to February 12, 2020, were enrolled in the study. West Court of Union Hospital, located in Wuhan, Hubei Province, the epidemic areas of COVID-19, is one of the major tertiary teaching hospitals, and has been mainly responsible for the treatments of COVID-19 patients assigned by the government. Professor Pinhua Pan, from Department of Respiratory Medicine in Xiangya Hospital, Central South University, was assigned as the director of National Medical Team of aiding Hubei to guide the clinical work for the treatment of COVID-19 in Union Hospital of Huazhong University of Science and Technology, Wuhan. All patients with COVID-19 enrolled in this study were diagnosed according to World Health Organization interim guidance [10]. This study was approved by the institutional ethics board of Union Hospital of Huazhong University of Science and Technology and ethics board of Xiangya Hospital, Central South University (No.202003049). Written informed consent was waived by the Ethics Commission of the designated hospital for emerging infectious diseases.

### Measurements

Demographic, clinical features, laboratory, and radiological findings, treatment strategy, and short-term prognosis data of the patients were obtained from their medical records. Clinical outcomes were followed up to March 12, 2020. All the data were checked by two senior physicians (P.P and J.Z). All patients enrolled to this study were laboratory-confirmed COVID-19 patients, and the diagnostic criteria of COVID-19 was based on the positive detection of viral nuclear acids. The severity of COVID-19 was defined based on the diagnostic and treatment guideline (Version 5-7) by the National Health Committee of China. Severe subtype was defined if a patient met one of the following criteria: 1) Respiratory distress with respiratory frequency ≥ 30/min; 2) Pulse oximeter oxygen saturation ≤ 93% at rest; 3) Oxygenation index (artery partial pressure of oxygen/inspired oxygen fraction, PaO2/FiO2) ≤ 300 mmHg. Critically ill subtype followed above criteria, and must meet one of the following criteria: 1) Needs mechanical ventilation because of respiratory failure; 2) Shock; 3) Combined with multiple organ failure that needs to be admitted to intensive care unit (ICU).

Pharyngeal swab specimens were collected from each patients for viral nucleic acid detection of SARS-CoV-2 using a real-time reverse-transcriptase polymerase-chain-reaction (RT-PCR) assay as previously described [11]. The viral nucleic acid testing were carried out by the clinical laboratory of Union Hospital of Huazhong University of Science and Technology in Wuhan. Laboratory indicators on admission, included the numbers of leucocytes, neutrophils, lymphocytes, percentages of lymphocyte, eosinophils, concentrations of C-reactive protein (CRP), procalcitonin (PCT), fasting blood glucose (FBG), lactate dehydrogenase (LDH), serum creatine kinase (CREA), blood urea nitrogen (BUN), creatine kinase isoenzyme MB (CK-MB), D-dimer, international normalized ratio (INR), prothrombin time (PT), activated partial thromboplastin time (APTT), and thrombin time (TT) were determined for each patient. All medical laboratory data were generated by the clinical laboratory of Union Hospital of Huazhong University of Science and Technology.

All data were reviewed and collected onto the standardized forms from the electronic medical records in the hospital, including case report forms, nursing records, laboratory and radiological findings were reviewed. Two senior physicians (P.P and J.Z) independently reviewed the data. Information on demographic data, symptoms, preexisting chronic comorbidities, computed tomographic images of chest, laboratory results were collected. All treatment strategy were collected during the hospitalization, such as antiviral therapy, antibiotic therapy, use of corticosteroid, and respiratory support. The time from illness onset to hospital admission were also recorded.

### Definition of conditions

Patients were identified as having diabetes if there was a documented medical history. Cardiovascular disease included coronary artery disease, congestive heart failure, or a history of myocardial infarction; reports of isolated hypertension were not included. Chronic pulmonary disease included chronic obstructive pulmonary disease (COPD), allergic airway disease or the use of supplemental oxygen at home. The presence of ARDS was designated by The berlin definition [12]. Cardiac injury was identified when the serum level of hypersensitive cardiac troponin I (hsTNI) was above the upper limit of the normal range (>28 pg/mL) or new abnormalities shown in electrocardiography and echocardiography [13]. Acute kidney injury (AKI) was classified on the basis of the highest serum creatinine level or urine output criteria based on KDIGO clinical practice guideline for AKI [14]. Prognosis was defined as discharged from hospital, not discharged yet, and death during hospitalization. The followed-up observation was conducted before March 12, 2020.

### Statistical Analysis

Continuous variables were shown as median and interquartile range (IQR), and compared by Mann-Whitney test since most laboratory data were with skewed distribution. Categorical variables were presented as counts and proportions, and compared by Chi-square test or Fisher’s exact test. The Cox proportional hazard model was used to determine the associations of diabetes and FBG with fatality of COVID-19, adjusting for potential confounders. Adjusted hazard ratio (aHR) with 95% confidence interval (CI) were presented as the effect size. All statistical analyses and graphs were generated and plotted using the GraphPad Prism version 7.00 software (GraphPad Software Inc) or SPSS version 25.0 (IBM, United States). A *P* value <0.05 was considered statistically significant.

## Results

### Baseline characteristics of the patients of COVID-19

A total of 258 consecutive laboratory-confirmed patients with SARS-CoV-2 infection were included and analyzed in the study, and 24% of them had diabetes. Demographic and clinical characteristics of the patients on admission were summarized by diabetes in **Table 1**. The median age was 64 years (IQR, 56-70; range, 23-92 years), and 138 (53.5%) were male. The median durations from illness onset to hospital admission were 12 days (IQR, 7-15). The most common symptoms at the illness onset were fever (212 [82.2%]), dry cough (173[67.1%]), polypnea (124 [48.1%]), and fatigue (98 [38%]). One hundred seventy-four patients (67.4%) had other preexisting chronic comorbidities, including hypertension (37.3%), cardiovascular disease (14.9%), cerebrovascular disease (4.4%), chronic pulmonary disease (3.6%), chronic kidney disease (3.69.2%), chronic liver disease (1.2%), and cancer (4.4%). There were no significant differences between patients with and without diabetes, with respect to age, sex, days of illness before hospital admission, and clinical signs and symptoms. Compared with those without diabetes, diabetic patients were more likely to have comorbidities of cardiovascular disease (23.8% vs. 12.3%, P=0.027) and chronic kidney disease (8.8% vs. 2.1%, *P*=0.027).

**Table 1.**
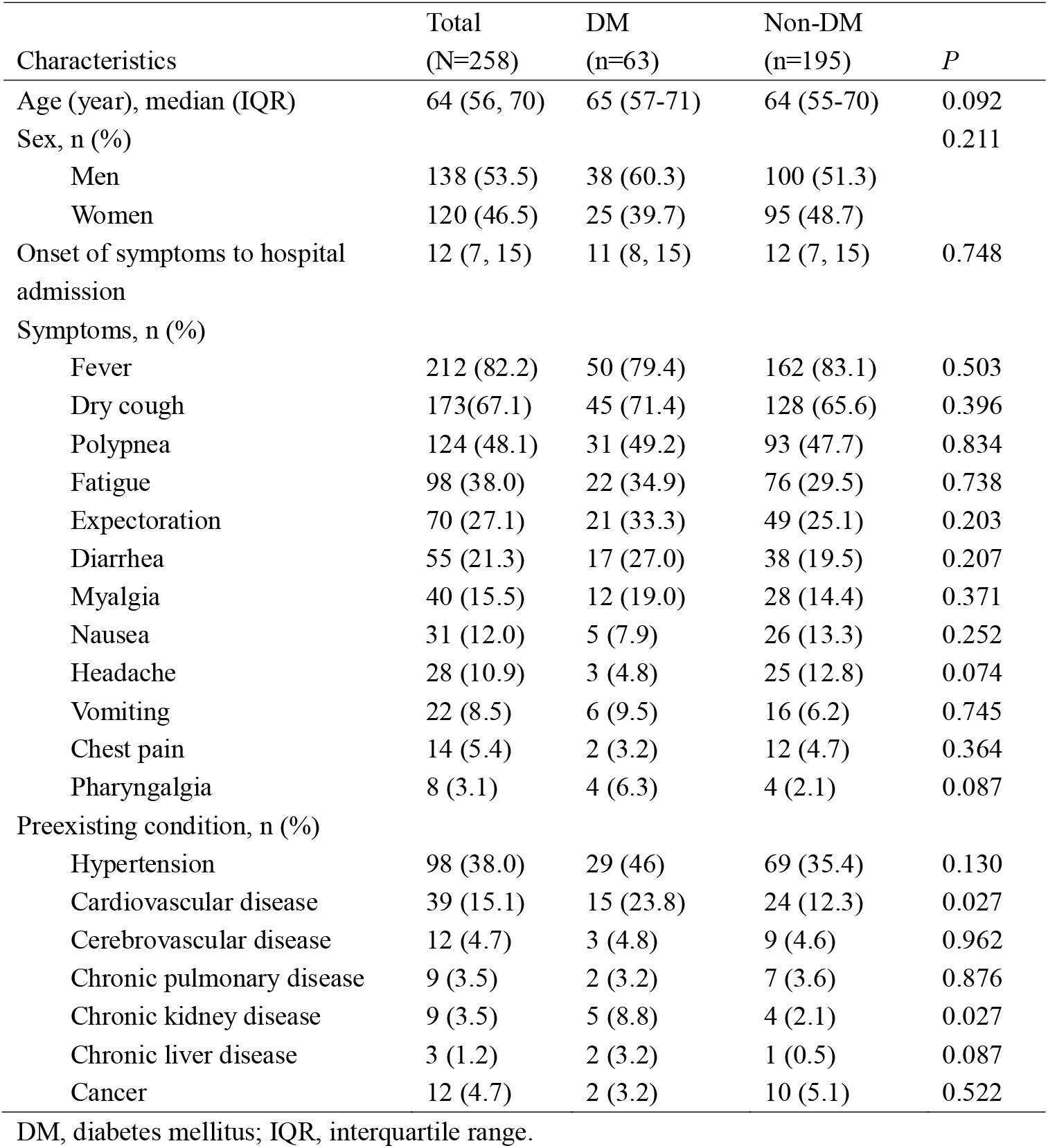
Characteristics of the patients with COVID-19.

### Laboratory and radiological findings in patients of COVID-19 on admission

The laboratory and radiological findings on admission of the COVID-19 patients with or without diabetes were presented in **Table 2**. The blood cell tests showed that most patients (81.0%) had normal leukocytes on the hospital admission, while 10.1% had increased leucocytes numbers and 8.9% had decreased numbers. However, the diabetic COVID-19 patients had a higher median leucocytes number (6.34[IQR: 4.66, 8.15] vs. 5.45[IQR: 4.31, 7.19], *P*=0.039) and median neutrophils numbers (4.49[IQR: 3.12, 6.91] vs. 3.82[IQR: 2.81, 5.39], *P*=0.022) compared with non-diabetic patients. COVID-19 patients with diabetes had more leucocytes increase (20.6% vs. 6.7%) but less leucocytes decrease (4.8% vs. 10.3%, *P*=0.004) than those patients without DM. The neutrophil-to-lymphocytes ratio (NLR) was significantly higher in diabetic patients compared to those without diabetes (median: 4.56[IQR: 2.69, 9.51] vs. median: 3.8[IQR: 2.25, 6.31], *P*=0.043). Interestingly, decreased eosinophil counts were also common in these patients (43%), but no significant difference in eosinophils counts and the ratio of patients with decreased eosinophil counts was found in patients with or without DM. Besides, a positive correlation between eosinophil and lymphocytes numbers on admission was observed (**Figure S1**), which was consistent with previous study [15]. Other laboratory findings showed no significant differences between the two groups of patients with respect to serum levels of CRP, PCT, and LDH, but diabetic COVID-19 patients had higher levels of FBG (median: 7.54[IQR: 6.37, 10.62] vs. median: 5.81[IQR: 5.32, 6.59], *P*<0.001), CREA (median: 74[IQR: 64.25, 95.78] vs. median: 67.8[IQR: 57.2, 78.23], *P*=0.005), BUN (median: 5.9[IQR: 4.09, 8.62] vs. median: 4.41[IQR: 3.58, 5.57], *P*<0.001), and CK-MB (median: 14[IQR: 10, 17] vs. median: 11[IQR: 9, 14], *P*=0.042) than non-diabetic patients. For coagulation function markers, patients with diabetes showed a slightly longer of TT (median: 16.1[15.25, 16.65] vs. median: 15.5[IQR: 14.9, 16.23], *P*=0.035) than those patients without diabetes. The radiological CT images showed the majority of patients (99.6%) had abnormal results with bilateral lesions.

**Table 2.** Laboratory and radiological findings of COVID-19 patients at admission.

| Variable | Total<br>(N=258) | DM<br>(n=63) | Non-DM<br>(n=195) | *P* |
| --- | --- | --- | --- | --- |
| WBC ($\times 10^9/L$), median (IQR) | 5.64 (4.37, 7.39) | 6.34 (4.66, 8.15) | 5.45 (4.31, 7.19) | 0.039 |
| < $3.5 \times 10^9/L$, n (%) | 26 (10.1%) | 13 (20.6%) | 13 (6.7%) | 0.004 |
| 3.5-9.5 $\times 10^9/L$, n (%) | 209 (81.0%) | 47 (74.6%) | 162 (83.1%) | |
| > $9.5 \times 10^9/L$, n (%) | 23 (8.9%) | 3 (4.8%) | 20 (10.3%) | |
| Neutrophil ($\times 10^9/L$), median (IQR) | 3.92 (2.85, 5.83) | 4.49 (3.12, 6.91) | 3.82 (2.81, 5.39) | 0.022 |
| Lymphocyte ($\times 10^9/L$), median (IQR) | 1.01 (0.71, 1.38) | 1.1 (0.65, 1.41) | 0.995 (0.72, 1.37) | 0.655 |
| < $1.1 \times 10^9/L$, n (%) | 146 (56.6%) | 30 (47.6%) | 116 (59.5%) | 0.099 |
| Lymphocyte percentage (%), median (IQR) | 18.4 (11.63, 26.08) | 16.4 (9.05, 25.1) | 18.65 (12.3, 26.7) | 0.118 |
| <20%, n (%) | 150 (58.1%) | 41 (65.1%) | 109 (55.9%) | 0.199 |
| NLR, median (IQR) | 3.95 (2.47, 7.07) | 4.56 (2.69, 9.51) | 3.8 (2.25, 6.31) | 0.043 |
| Eosinophil ($\times 10^9/L$), median (IQR) | 0.03 (0, 0.07) | 0.02 (0, 0.08) | 0.03 (0, 0.07) | 0.553 |
| CRP (mg/L), median (IQR) | 30.75 (4.94, 69.17) | 30.75 (4.53, 81.72) | 30.68 (5.75, 67.37) | 0.765 |
| PCT (ng/ml), median (IQR) | 0.07 (0.05, 0.14) | 0.07 (0.05, 0.26) | 0.07 (0.04, 0.13) | 0.116 |
| LDH (U/L), median (IQR) | 268.5 (206, 355.75) | 318 (195.5, 426.3) | 264.5 (207, 338) | 0.205 |
| FBG (mmol/L), median (IQR) | 6.04 (5.41, 7.20) | 7.54 (6.37, 10.62) | 5.81 (5.32, 6.59) | <0.001 |
| CREA ($\mu\text{mol/L}$), median (IQR) | 69.6 (57.93-79.9) | 74 (64.25, 95.78) | 67.8 (57.2, 78.23) | 0.005 |
| BUN (mmol/L), median (IQR) | 4.65 (3.66, 6.34) | 5.9 (4.09, 8.62) | 4.41 (3.58, 5.57) | <0.001 |
| CK-MB (U/L), median (IQR) | 12 (9, 15) | 14 (10, 17) | 11 (9, 14) | 0.042 |
| D-Dimer ($\mu\text{g/mL}$), median (IQR) | 0.56 (0.27, 1.62) | 0.87 (0.35, 2.46) | 0.54 (0.25, 1.51) | 0.046 |
| PT (sec), median (IQR) | 13 (12.3, 13.9) | 12.05 (12.8, 14) | 13 (12.4, 13.9) | 0.394 |
| APTT (sec), median (IQR) | 36.75 (33.45, 40.65) | 37 (32.55, 42.55) | 36.6 (33.65, 40.5) | 0.760 |
| TT (sec), median (IQR) | 15.6 (14.93, 16.3) | 16.1 (15.25, 16.65) | 15.5 (14.9, 16.23) | 0.035 |
| INR, median (IQR) | 1 (0.94, 1.09) | 0.98 (0.91, 1.1) | 1 (0.94, 1.09) | 0.326 |
| Abnormal Chest CT images, n (%) | 254 (98.4%) | 62 (98.4%) | 192 (98.5%) | 0.978 |
| Bilateral lung involved | 247 (95.7%) | 60 (95.2%) | 187 (95.9%) |  |
| Left lung involved only | 3 (1.2%) | 1 (1.6%) | 2 (1.0%) |  |
| Right lung involved only | 4 (1.6%) | 1 (1.6%) | 3 (1.5%) |  |

DM, diabetes mellitus; IQR, interquartile range; WBC, white blood cell; NLR, neutrophil-to-lymphocyte ratio; CRP, C-reactive protein; PCT, procalcitonin; LDH, lactate dehydrogenase; FBG, fasting blood glucose; CREA, serum creatine; BUN, blood Urea nitrogen; CK-MB, creatine kinase isoenzyme MB; PT, prothrombin time; APTT, activated partial thromboplastin time; TT, thrombin time; INR, international normalized ratio.

**Figure S1**. Correlations between blood lymphocytes and eosinophils on admission.

### Analysis of severity, treatment and prognosis of patients with COVID-19

Next, we compared the severity, treatment, and short-term prognosis of the COVID-19 patients with and without diabetes in **Table 3**. Compared with non-diabetes subjects, diabetic patients were more likely to develop severely or critically ill subtypes (*P*=0.028) with more complications including acute respiratory distress (38.1% vs. 19.5%, *P*=0.001), acute cardiac injury (14.5% vs. 5.1%, *P*=0.016), and had more antibiotic therapy (74.6% vs. 59.0%, *P*=0.026), non-invasive and invasive mechanical ventilation (*P*=0.037). As of March 12, 2020, only 33.7% patients were discharged from the hospital. Patients with diabetes had a higher fatality rate than those without diabetes (11.1% vs. 4.1%, *P*=0.039).

**Table 3.** Disease severity, treatment, and prognosis of COVID-19 patients.

| Variable | Total<br>(N=258) | DM<br>(n=63) | Non-DM<br>(n=195) | *P* |
| --- | --- | --- | --- | --- |
| Severity, n (%) |  |  |  |  |
| Mild to moderate | 87 (33.7) | 18 (28.6) | 69 (35.4) | 0.028 |
| Severe | 116 (45.0) | 24 (38.1) | 92 (47.2) |  |
| Critical | 55 (21.3) | 21 (33.3) | 34 (17.4) |  |
| Complications, n (%) |  |  |  |  |
| Acute respiratory distress | 62 (24.0) | 24 (38.1) | 38 (19.5) | 0.001 |
| Acute cardiac injury | 19 (7.4) | 9 (14.5) | 10 (5.1) | 0.016 |
| Acute kidney injury | 7 (2.7) | 3 (4.8) | 4 (2.1) | 0.250 |
| Medication, n (%) |  |  |  |  |
| Antiviral agent | 246 (95.3) | 60 (95.2) | 186 (95.4) | 0.962 |
| Antibiotic | 162 (62.8) | 47 (74.6) | 115 (59.0) | 0.026 |
| Corticosteroid | 74 (28.7) | 17 (27.0) | 57 (29.2) | 0.792 |
| Oxygen support, n (%) |  |  |  |  |
| Nasal cannula | 148 (57.4) | 30 (47.6) | 118 (60.5) | 0.037 |
| High-flow oxygen | 24 (12.4) | 8 (12.7) | 24 (12.3) |  |
| Non-invasive ventilation | 26 (10.1) | 10 (15.9) | 16 (8.2) |  |
| Invasive mechanical ventilation | 16 (6.2) | 7 (11.1) | 9 (4.6) |  |
| ECMO | 1 (0.4) | 1 (1.6) | 0 (0) |  |
| Prognosis, n (%) |  |  |  |  |
| Discharged | 87 (33.7) | 16 (35.7) | 71 (36.4) | 0.039 |
| Not discharged yet | 156 (60.5) | 40 (63.5) | 116 (59.5) |  |
| Death | 15 (5.8) | 7 (11.1) | 8 (4.1) |  |

DM, diabetes mellitus; ECMO, extracorporeal membrane oxygenation.

### Analysis of associations of diabetes and FBG with death in COVID-19 patients

To further assess the association of diabetes and FBG with the fatality of COVID-19, Cox proportional hazard model was carried out, and the results (**Table 4**) showed that comorbid diabetes was an independent risk factor for death in COVID-19 patients, after adjusting for age (aHR=2.804; 95% CI: 1.008, 7.797; *P*=0.048) or additionally adjusted for the cardiovascular diseases and chronic kidney diseases (aHR =2.840; 95% CI: 1.007, 8.007; *P*=0.048) or additionally adjusting for laboratory markers (aHR=3.641; 95% CI: 1.086, 12.214; *P*=0.036). We also found that higher FBG level on admission was an independent predictor for death in COVID patients as well, after adjusting for the aforementioned covariates (AHR=1.187, 95% CI: 1.078, 1.306; *P*<0.001).

**Table 4.**
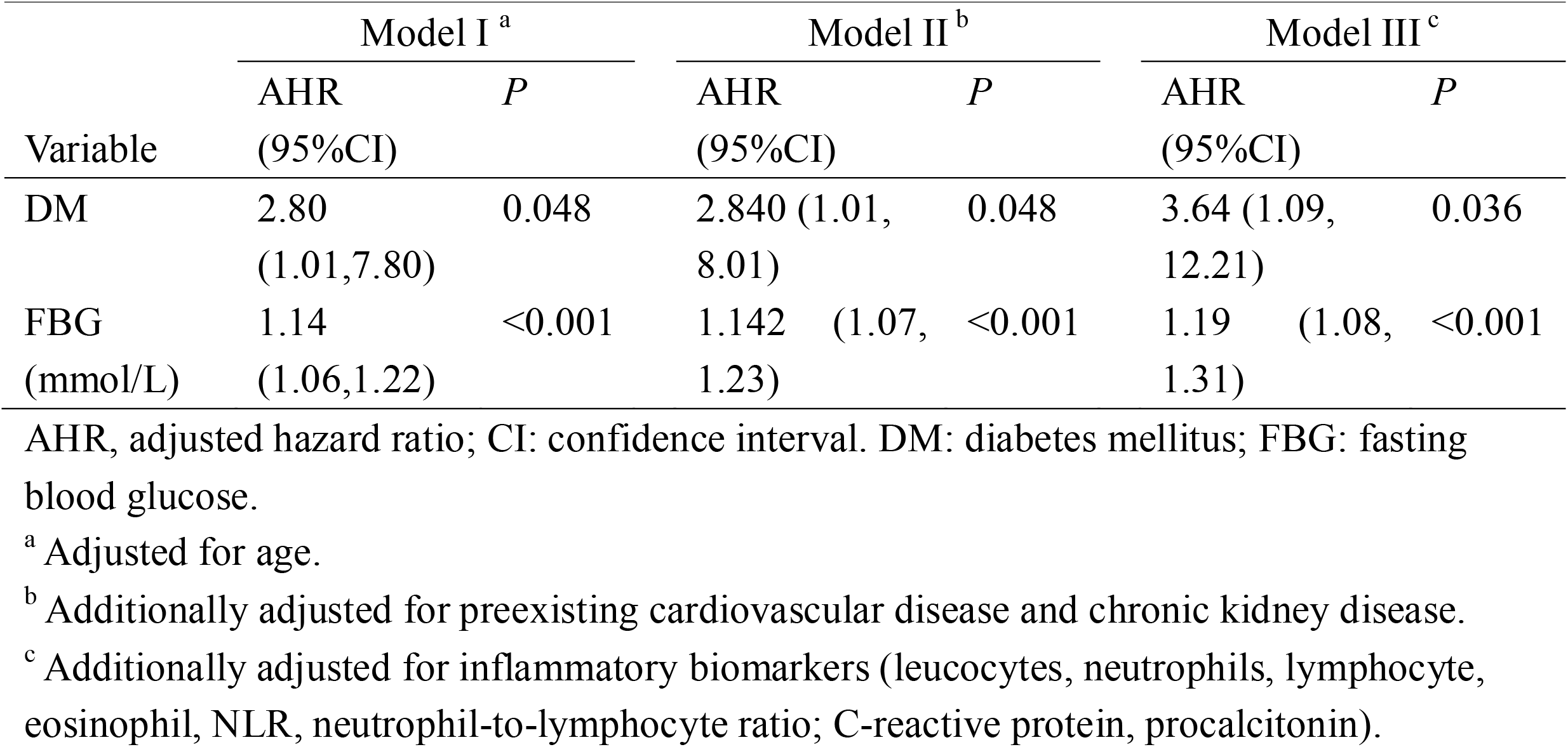
Associations of diabetes and FBG with fatality of COVID-19 in Cox proportion hazard models.

| Variable | Model I <sup>a</sup> |  | Model II <sup>b</sup> |  | Model III <sup>c</sup> |  |
| --- | --- | --- | --- | --- | --- | --- |
|  | AHR<br>(95%CI) | *P* | AHR<br>(95%CI) | *P* | AHR<br>(95%CI) | *P* |
| DM | 2.80<br>(1.01,7.80) | 0.048 | 2.840 (1.01,<br>8.01) | 0.048 | 3.64 (1.09,<br>12.21) | 0.036 |
| FBG<br>(mmol/L) | 1.14<br>(1.06,1.22) | <0.001 | 1.142 (1.07,<br>1.23) | <0.001 | 1.19 (1.08,<br>1.31) | <0.001 |

AHR, adjusted hazard ratio; CI: confidence interval. DM: diabetes mellitus; FBG: fasting blood glucose.
<sup>a</sup> Adjusted for age.
<sup>b</sup> Additionally adjusted for preexisting cardiovascular disease and chronic kidney disease.
<sup>c</sup> Additionally adjusted for inflammatory biomarkers (leucocytes, neutrophils, lymphocyte, eosinophil, NLR, neutrophil-to-lymphocyte ratio; C-reactive protein, procalcitonin).

## DISCUSSION

In this retrospective cohort study, we characterized 258 COVID-19 patients with respect to demographics, clinical features, preexisting chronic comorbidities, treatment, and short-term prognosis. We found that COVID-19 patients had a relatively high proportion (24%) of diabetes, and demonstrated that diabetes was associated with alterations in laboratory markers, severer clinical subtype at presentation, and poorer prognosis compared to those without diabetes, after SARS-CoV-2 infection.

To the best of our knowledge, the study was the first to investigate the clinical characteristics and prognosis of COVID-19 patients with diabetes. The prevalence of diabetes mellitus is sharply climbing in China during the past decades. According to the latest nationally representative cross-sectional survey in mainland China in 2013, the estimated prevalence of diabetes in elderly participants (≥60 years old) was 20.2% [4]. Previous studies reported 9% to 14% prevalence of diabetes in COVID-19 patients [1, 8, 13, 15]. Here, we reported a higher prevalence rate of diabetes in these patients, which might be due to the larger proportion of geriatric patients infected by SARS-CoV-2 in our study. The median age of all the participants was 64 years old, which was older than the data previously reported [1, 7, 8]. In the current study, 53.5% of the patients were male, and the percentage is similar to that reported by Wang et al [8] and Zhang et al [15]. Besides diabetes mellitus, hypertension (38.0%) and cardiovascular diseases (15.1%) were also common underlying chronic illness, and diabetic COVID-19 patients seemed to have more comorbidities of cardiovascular diseases and chronic kidney diseases in the current study.

The laboratory findings on admission showed that leucocytes and neutrophils counts and the proportion of increased leucocytes were higher in COVID-19 patients with diabetes than those without, which might be explained by the fact that diabetic patients were more susceptible to pathogens after viral infection due to lower immune function. During hospitalization, the diabetic patients were more likely to receive antibiotic therapy as well. Decreased lymphocytes counts and eosinophil counts were also common in these patients; this was consistent with the results of previous studies [15]. However, no significant differences were found in the cell counts and percentage lymphocytes and eosinophil counts between COVID-19 patients with or without diabetes. The data revealed that diabetic COVID-19 patients had a higher NLR, which was recently reported as a predictor of severity of COVID-19 in the early stage [16]. We found that COVID-19 patients with diabetes were more likely to develop severely or critically ill subtypes, including more complications with ARDS, acute cardiac injury, resulting in receiving more antibiotic therapy and mechanical ventilation. Cox regression model indicated that both diabetes and FBG level on admission were independent predictor for the fatality of COVID, after adjusting for potential confounders. Based on these findings, we deliberately concluded that diabetes was associated with deteriorated disease severity and poorer prognosis in patients with COVID-19.

This is the first report to demonstrated that the diabetes was associated with greater disease severity and poorer prognosis in COVID-19 patients. An increasing number of studies have shown that diabetic patients had higher mortality and morbidity of severe medical illness, such as myocardial infarction, and high FBG plays an independent predictive role in hospitalized non-diabetic patients as well [17, 18, 19]. Diabetes has also been identified as a significant risk factor for severe disease following respiratory tract infections [20]. Several studies demonstrated that diabetes was associated with increased the risks of severity and mortality after SARS-CoV and MERS-CoV infection [21, 22, 23, 24], and FBG level was an independent predictor for fatality in patients with SARS [21].

Additionally, we also found diabetic COVID-19 patients had more preexisting cardiovascular disease, and were more susceptible to have acute cardiac injury during hospitalization, which might increase the possibility of short-term poor prognosis in diabetic patients after SARS-CoV-2 infection. Previous study reported that diabetic patients who received intensive glycemic control had lower risk of cardiovascular events [25]. Nevertheless, we could deliberately conclude that diabetes and FBG were independent predictive risks for poor outcomes in COVID-19 patients after adjusting those cofounders and mediators.

Diabetes results in a proinflammatory homeostatic immune response skewed toward helper T cell 1 (Th1) and T17 cells and a decrease in regulatory T cells (Treg) [26]. Immune dysfunction of diabetes alone or following infection has been reported for a wide variety of immune cells, not just macrophages, monocytes and CD4^+^ T cells [26]. A recent study reported the number of total T cells, CD4^+^ and CD8^+^T cell subsets were substantially reduced and functionally exhausted in COVID-19 patients, especially among geriatric and critically ill patients who required ICU admission [27]. Kulcsar KA *et al* showed that diabetic mice presented a prolonged phase of severe disease and delayed recovery after MERS-CoV infection, which was attributed to dysregulated immune response with lower inflammatory monocytes/macrophages and CD4^+^ T cells [28]. Thus, optimal management of diabetes and intensive glycemic control may help prevent the occurrence of life-threatening infections and complications associated with diabetes mellitus, and the increased susceptibility of infections due to impaired cellular and humoral immunity.

## Limitations

Several limitations should not be ignored in the study. First, this study was a retrospective study, with the fact that we included a very small proportion of patients with laboratory-confirmed SARS-CoV-2 infection in Wuhan. Berkson bias might be introduced since asymptomatic patients and those with mild symptoms were less likely to be enrolled. Second, due to the massive number of patients and the lack of medical resources, the interval from the illness onset to hospital admission was more than 10 days for most patients, which could further complicate and deteriorate the illness, with different extent. Nevertheless, patients with diabetes had similar interval from onset to admission compared with those without diabetes. Third, at the time of the study submission, most of patients were not discharged yet, and the final survival outcome could not be determined now, and the long-term prognosis were not observed.

## Conclusions

In the current study, we demonstrated that diabetes mellitus is associated with greater disease severity and poorer short-term outcomes including death. Stronger personal prophylactic strategies are advised for diabetic patients, and more intensive surveillance and treatment should be considered when they are infected with SARS-CoV-2, especially in geriatric patients or those had preexisting comorbidities.

## Data Availability

Data is available upon request to corresponding authors.

## Acknowledgement

We thank all the patients involved in the study. We thank all medical staff who are fighting against this public crisis.

